# Validating a novel driving simulation-based MWT against the standard MWT in an OSA-cohort challenged by CPAP-withdrawal (DS-MWT2) – Protocol for a monocentric, controlled, randomized, crossover trial

**DOI:** 10.64898/2026.01.18.26344362

**Authors:** Veronika Gambin, Nan Li, Esther Irene Schwarz, Kristina Keller, Stefan Lakämper

## Abstract

**Background:** Excessive daytime sleepiness (EDS) is a major yet under-recognized contributor to road traffic accidents. Traditional diagnostic tools, such as the Maintenance of Wakefulness Test (MWT), assess wakefulness under passive conditions but may not accurately reflect real-world driving risks. To address this gap, we have piloted a Driving Simulation–based MWT (DS-MWT), designed to evaluate sleepiness in an ecologically valid driving scenario. The present study aims to validate the novel DS-MWT against the classical MWT in a functionally relevant cohort of patients with obstructive sleep apnoea (OSA).

**Methods:** The present monocentric, randomized, controlled, within-subject crossover trial will include 54 participants: 36 patients with OSA undergoing *≥* 7-day CPAP withdrawal (W) or continuation (C), and 18 healthy controls. The study employs a well-established CPAP-withdrawal model in patients with prior optimal treatment adherence to transiently induce EDS under controlled conditions. A healthy control group is included to enable between-group comparisons. The primary expected outcome is the difference in mean latencies between MWT and DS-MWT, determined during four standardized test sessions per condition. Secondary exploratory outcomes are defined as the presence, direction, and magnitude of differences or correlations between treatment status (CPAP withdrawal vs. continuation) and driving performance metrics (e.g., lateral position, speed, lane departures, etc.), EEG and eye-tracking features, subjective sleepiness scores, at-home polysomnography (PSG) parameters, and metabolomic biomarkers (saliva, exhaled breath and dried blood spots). Data will be analyzed using linear mixed models, repeated-measures ANOVA, and predictive modeling with cross-validation.

**Discussion:** This trial addresses a critical limitation in sleep and traffic medicine by introducing a realistic, supposedly more ecologically valid alternative to standard sleepiness assessment tools. The DS-MWT may enhance clinical decision-making regarding fitness to drive (FTD) and provide a framework for identifying physiological and behavioral markers of sleepiness in realistic conditions.

**Trial registration:** ClinicalTrials.gov Identifier: NCT06872593, released on 12.03.2025,

https://clinicaltrials.gov/study/NCT06872593

Swiss National Clinical Trial Portal SNCTP000006301, released on 19.03.2025,

https://www.humanforschung-schweiz.ch/en/trial-search/study-detail/66469

## Introduction

### Background and rationale

Sleepiness at the wheel – frequently termed drowsy driving - is one of the major contributors to road accidents [1] with increasing need for an adequate legal regulation [2, 3]. Recent work foregrounds the necessity for appropriate tools to assess sleepiness behind the wheel [4]. The proposed study aims to validate a recently developed naturalistic driving simulation-based test [5], which seeks to quantify the extent of sleepiness during a standardized monotonous night-time driving task.

In medical terms, sleepiness is often referred to as excessive daytime sleepiness (EDS), a symptomatic condition characterized by an increased propensity to fall asleep during waking hours. EDS may result from insufficient or compromised sleep, caused by psychosocial factors (e.g., stress), voluntary sleep dept, shift work, or medical conditions such as narcolepsy or obstructive sleep apnoea (OSA) [6]. Among the medical causes of EDS, OSA is one of the most prevalent and well-characterized, it is marked by recurrent episodes of upper airway obstruction during sleep, resulting in fragmented sleep and intermittent hypoxia [7]. Although the primary treatment of OSA with continuous positive airway pressure (CPAP) can alleviate EDS [8], a substantial subset of patients continues to report EDS despite receiving an optimized therapy [9]. Untreated EDS poses a substantial risk to traffic safety and is considered one of the highest-ranking causes of motor vehicle accidents [1, 10, 11].

Sleepiness and its dangers at the wheel might subjectively not be registered by the affected drivers [12], where the subjective perception of sleepiness frequently fails to align with objective measures of wakefulness [13].

Given the limitations of subjective perception, objective tools are essential for the assessment of sleepiness. There exists a variety of partially complementary tools to evaluate the extent of EDS. Mean sleep latencies obtained in the maintenance of wakefulness tests (MWT) are commonly used in assessing EDS. MWT latency refers to the time it takes for a person to fall asleep during a 40-minute-long sitting session. It is defined as the interval (in minutes) from the start of the session (lights out) until the start of the first epoch of any stage of sleep (an epoch of N1, N2, N3, or R) [14].

However, there is inconsistent or insufficient evidence for MWT to reliably predict the fitness to drive (FTD) in general, potentially due to the test’s susceptibility to individual motivational factors [15]. Moreover, the cost and time demands of MWTs limit their feasibility for widespread application in driving license certification [16].

Currently debated is whether the MWT should be replaced or supplemented by emerging tools [4], such as future roadside metabolomic tests for detecting sleepiness in traffic [17, 18], or whether the MWT itself should be adapted to more effectively communicate the risks of EDS for road safety and its relevance in determining FTD [19].

In line with these arguments, a driving simulation-based MWT (DS-MWT) was developed [5]. Specifically, the operational structure of MWT was retained, including electroencephalography (EEG) and video monitoring across four 40-minute soporific sessions. The drowsiness-inducing conditions were achieved through a highly monotonous, nighttime car-following scenario, which directly mimics real-life driving environments in which a considerable proportion of vehicle accidents occur [20]. In addition to the solely EEG-based criteria used to define the latency in MWT, the DS-MWT latency was complemented by a “naturalistic” criterion, an accident with closed eyes (“sleep accident,” SA). The pilot resulted in about 23% of driving sessions (runs) terminating with SA in the unstructured cohort, yielding a run latency, and thus inherently displayed sufficiently soporific conditions of DS-MWT [5]. Participant feedback indicated high user acceptance and increased ecological face validity.

While the pilot served as a successful feasibility study, offering proof of concept with encouraging outcomes, these findings warrant a systematic comparison of MWT and DS-MWT based on their resulting latencies. It also remains to be determined whether these latencies reliably reflect clinical parameters of EDS associated with underlying medical conditions such as OSA. Assessing the extent of EDS in the OSA-affected population may be further complicated by suboptimal CPAP adherence in CPAP-naive patients, potentially leading to inconsistent and inconclusive results. A previously established experimental model suggests overcoming these drawbacks by employing a restricted pool of patients effectively treated and compliant with CPAP-therapy [21, 22]. Short-term CPAP withdrawal in patients with previously optimal adherence has been shown to reactivate OSA and induce associated consequences, such as a deterioration of daytime symptoms and psychomotor performance [22, 23]. However, it remains unclear whether EDS itself is consistently and measurably reactivated in this model, as no systematic data on its recurrence are currently available. Overall, this trial may contribute valuable insight into whether short-term CPAP withdrawal can reliably induce EDS and whether this effect can be captured through objective measures.

This constellation of unresolved questions constitutes an important evidence gap for both medical experts in pulmonology and traffic medicine, but also for affected drivers. This gap will be addressed through a systematic comparison of classical and simulation-based MWTs by means of their resulting latencies. In a within-subject design involving 36 effectively treated and highly CPAP-adherent OSA patients, latency variations will be analyzed in response to controlled CPAP therapy withdrawal (W) and continuation (C), each maintained for a period of at least 7 days. A healthy control group (N=18) is included to enable comparison of MWT- and DS-MWT- latencies across clinical and non-clinical populations.

### Objectives

The primary objective of this trial is to determine whether there is a statistically significant difference in latency between the classical MWT and the DS-MWT. The primary hypothesis assumes no statistically significant difference between MWT and DS-MWT latencies. This hypothesis will be tested exclusively using mean latency values, independent of secondary and exploratory analyses. Latency differences will be measured in relation to the intervention CPAP withdrawal (OSA-affected only) and in relation to the test sequence (both OSA-affected and healthy control groups).

Secondary, exploratory objectives are designed to evaluate the effects of the intervention (CPAP withdrawal vs. CPAP continuation) and to relate these effects to those observed in a healthy control group. The following secondary objectives are based on a range of behavioral, physiological, and performance-related parameters.

The study will investigate potential associations between CPAP treatment status and driving performance indicators, including the standard deviation of lateral position (SDLP), driving speed, following distance, and steering wheel angle variability. These driving parameters will also be analyzed in relation to subjective perception of sleepiness, measured by the Epworth Sleepiness Scale (ESS) and the Karolinska Sleepiness Scale (KSS).

The relationship between CPAP condition and eye-tracking parameters, including blink rate, blink duration, eye openness, gaze direction and duration, and attention distribution across predefined areas of interest (e.g., windshield, side mirrors, rear mirror), will also be explored. In parallel, EEG-derived features will be examined as supplementary indicators that may reflect sleepiness, including microsleep-like episodes (MSEs) and individual electrophysiological signatures of sleep–wake transitions.

In addition, based on at-home polysomnography (PSG) recordings preceding each MWT and DS-MWT, night-to-night variability in the sleep quality will be examined in both groups, as well as key OSA parameters in the patient group, such as the apnoea–hypopnea index (AHI), oxygen desaturation index (ODI) and cumulative time with oxygen saturation below 90% [24].

Finally, the potential utility of noninvasive biological samples, including saliva, exhaled breath, and dried blood spots, for identifying proteomic biomarkers associated with sleepiness will be explored [17, 18]. These biomarkers will be analyzed in relation to sleep quality, CPAP treatment condition and subjective sleepiness metrics in the OSA group, and will be compared to baseline values from healthy participants.

## Materials and methods

### Trial setting

This study will be conducted solely on the premises of the University of Zurich, Zurich, Switzerland. Data collection will be conducted at the Traffic Medicine Department of the Institute of Forensic Medicine, University of Zurich.

### Trial status

The trail is recruiting. The original study protocol was first approved in version 1.1 by the ethics committee and amended twice, with the current protocol version being 1.3 (dated 08/07/2025). The planned overall duration of the study is 24 months, with 01/05/2025 being First-Participant-in (FPFV, anticipated date of the screening/enrollment of the first patient) and 30/04/2027 being Last-Participant-Out (LPLV, anticipated date of the last participant’s final visit). A published manuscript or submission of the final report will be handed in by 30/04/2028.

### Trial design

This study is a monocentric, controlled, within-subjects, randomized, crossover clinical trial in a cohort of OSA patients and in a healthy control group. The study consists of two equivalent arms designed to assess the impact of the test sequence based on the initial condition. The protocol will be reported following the Standard Protocol Items: Recommendations for Interventional Trials (SPIRIT) guideline S1 File [25].

### Eligibility criteria

#### Inclusion criteria (OSA patients)

- adult driver

- diagnosed OSA

- established CPAP-treatment regime, highly adherent and compliant within the last 6 months (*>* 5h*, >* 80% of days)

- at impaired eyesight with more than +/- 5 diopter or astigmatism, contact lenses are required (for eye tracking (ET))

#### Inclusion criteria (healthy comparison group)

- adult driver

- no declared psychiatric disorders

- no declared sleep-related diagnosis

- at impaired eyesight with more than +/- 5 diopter or astigmatism, contact lenses are required (for ET)

#### Exclusion criteria (for both groups)

-sensibility to motion sickness (kinetosis, dizziness, etc. in 5 min screening drive)
- professional drivers (if working during the study period)
- inability to understand the study procedure for linguistic or cognitive reasons

Based on the inclusion criteria and the preselected pool for recruitment, no vulnerable participants, as defined by the Swiss HRA (chapter 3, i.e., children, adolescents, adults lacking capacity, or prisoners), will be included. However, participation in the experimental procedure does not pose any additional risks for pregnant women, nor for patients, healthy adults or the unborn child. Therefore, the participation of pregnant individuals is possible, and a pregnancy test is not required.

### Recruitment

The total sample size for the study is N_total_ = 54, comprising an intervention group of N_I_ = 36 patients with OSA and a healthy comparison group of N_H_ = 18. Details of the sample size calculation are presented in section Sample size calculation.

#### Intervention group (OSA patients)

Participants in the intervention group will be recruited from a restricted pool of patients treated at the Sleep Disorders Center and Pulmonary Division of the University Hospital Zurich. This population is characterized by high adherence to CPAP therapy and regular clinical follow-up. Eligible individuals are preselected by the partner institution based on clinical criteria, including the absence of substantial comorbidities (e.g., major affective, somatic, or neurological disorders), cognitive impairments, or increased risk associated with CPAP withdrawal.

Although gender balance is desired, it cannot be guaranteed in the intervention group due to the significantly higher prevalence of OSA in males. Given the limited sample size and inter-individual variability of outcome measures, retrospective stratification by gender is not planned at this stage.

#### Healthy comparison group

Healthy participants will be recruited from the general population. Recruitment will be facilitated through institutional communication channels of the University of Zurich, including digital platforms and email distribution. Additionally, advertisements will be placed in local schools and public areas throughout Zurich to inform about the open recruitment status.

#### Recruitment Process

First contact will be offered by a flyer containing contact information to the investigators. Interested individuals can reach out to the study team via phone, email, video call (optional), or in-person at the study site. Investigators will provide detailed information about the study objectives, design, procedures, duration, risks, benefits, and any potential discomforts. These explanations can be delivered individually or in groups, using the participant’s preferred communication method.

All participants will receive a comprehensive information sheet and informed consent form, ensuring that they have sufficient detail to make an informed decision regarding their participation. Enough time, as much as the participant desires, will be given for consideration, and all participants will be informed that participation is entirely voluntary, with the option to withdraw at any time without consequences for their ongoing medical care.

As part of the screening process, all prospective participants will complete a brief, five-minute simulated drive to assess for motion sickness (kinetosis or dizziness), which constitutes an exclusion criterion. This session also ensures that participants fully understand the simulation task requirements before enrollment.

#### Consent and Assent

Verbal and written informed consent will be obtained by the sponsor-investigator or his designee (certified by GCP) prior to any study-specific procedures, including the collection of biological samples. The informed consent process will be documented in the participant’s file. Any deviations from the consent process as outlined in the protocol will be recorded and justified.

The participant will receive a copy of the signed consent form; the original will be retained in the study files. Consent includes permission for data collection, storage, analysis, and verification by study monitors or the ethics committee. For any ancillary studies not explicitly covered in the primary study’s informed consent form, additional written informed consent will be obtained from each participant prior to their involvement.

### Intervention and comparator

The intervention consists of a *≥*7-day withdrawal of CPAP therapy in patients with established adherence to treatment.

To reliably induce EDS for assessment, the study employs a validated intervention model that involves temporarily withdrawing CPAP therapy in patients with OSA [22]. The intervention arm involves discontinuation of CPAP therapy for a minimum of seven consecutive days prior to the assessment period. The assessments are conducted after either one or two weeks on the respective treatment, depending on the assigned crossover sequence.

Discontinuation or modification of the allocated interventions is not anticipated under normal study conditions. However, discontinuation may occur in case of adverse events or if participants fail to comply with protocol requirements. Additionally, if necessitated by organizational constraints or participant-related circumstances, a switch between treatment conditions may be permitted.

Concomitant care is managed with attention to participant well-being. If a participant experiences discomfort or requires assistance, a designated contact person may be called to escort them. If no such person is available, transportation to the participant’s home will be arranged by taxi. In the case of adverse events (AEs) during or following the study, participants are instructed to contact the sponsor-investigator or study physician, who will initiate appropriate medical follow-up. At the discretion of the clinical team, additional follow-up visits may be scheduled as needed.

### Participant timeline

The overview of the participant timeline is shown in the SPIRIT [25] schedule of enrollment, interventions, and assessment Fig.1.

**Fig 1.**
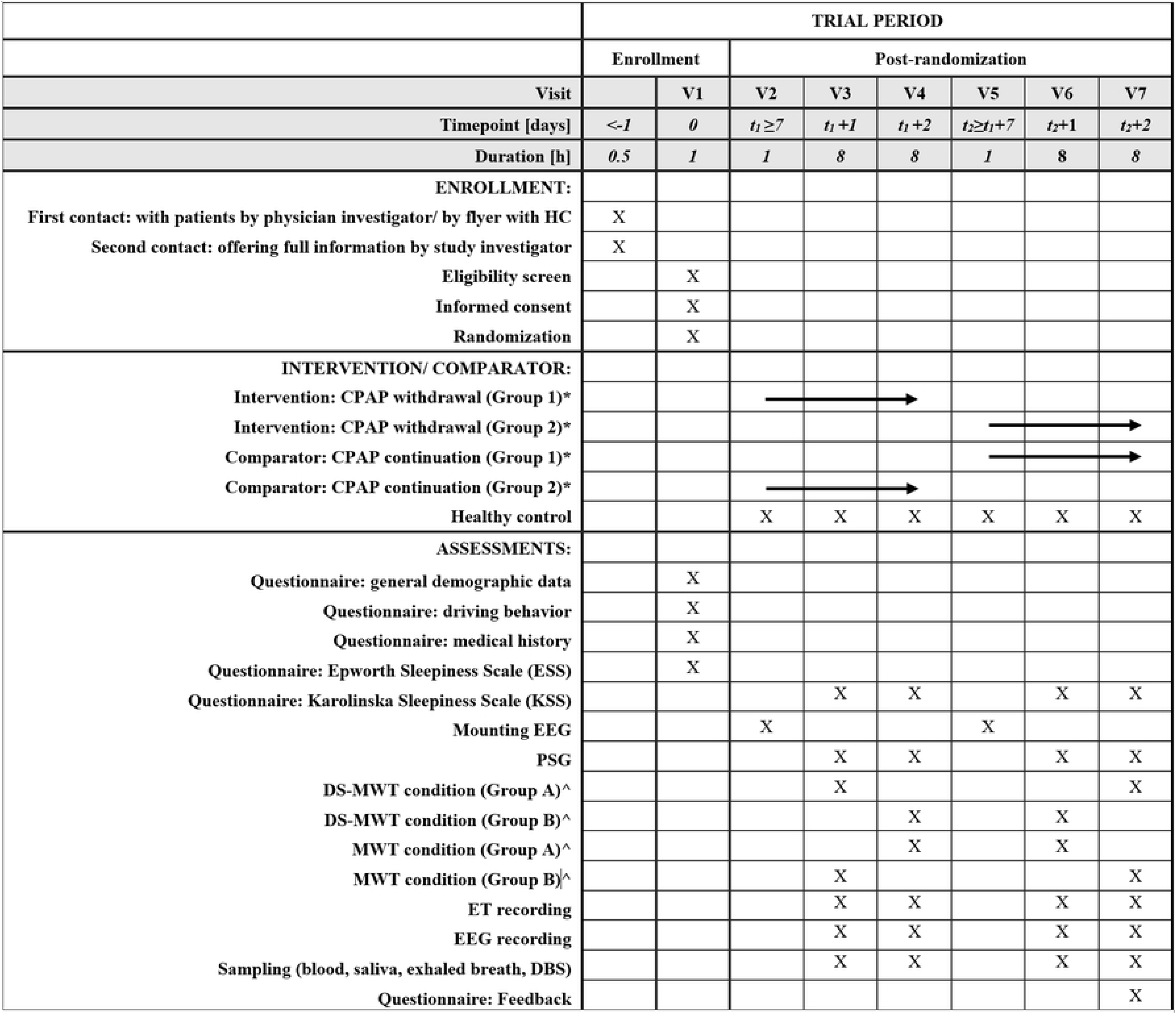
SPIRIT schedule of enrollment, interventions, and assessments. V = visit; HC = healthy control group; PSG = at-home polysomnography prior to respective visit; DS-MWT = driving simulation-based maintenance of wakefulness test; MWT = maintenance of wakefulness test; CPAP = continuous positive airway pressure; EEG = electroencephalography; ET = eye tracking; DBS = dried blood spots. Visit 1 comprises inclusion and a brief screening. Visit 2 and Visit 5 serve merely to mount PSGs. ∗ = prior to V2-V4 patients are randomly assigned to either a starting condition CPAP withdrawal (Group 1) or to a starting condition CPAP continuation (Group 2), and crossover to the respective other condition during V5-V7. ^ = participants are randomly assigned to either DS-MWT (Group A) or MWT (Group B) starting condition during V3, V4 and then crossover their condition during V6, V7.

The study comprises seven visits per participant, combining in-laboratory assessments with home-based PSG and interventional testing. Visit 1 involves participant onboarding, including informed consent, eligibility confirmation, a brief screening drive (to assess for motion sickness as an exclusion criterion), and randomization by lottery. This visit lasts approximately 1 hour.

Visits 2 and 5 are dedicated to mounting of EEG electrodes for home-based PSG recordings. Each of these visits requires approximately 1 hour of participant presence. PSGs are recorded overnight on the nights prior to visits 3, 4, and 6, 7.

Visits 3, 4, 6, and 7 include the primary experimental procedures: MWT (M) and the DS-MWT (D). These assessments comprise four 40-minute-long sessions (also termed “runs”) with EEG measurements including video, starting at 9 a.m., 11 a.m., 1 p.m. and 3 p.m.

Each session is preceded by a subjective sleepiness rating using KSS questionnaire. During the MWT, participants sit quietly with eyes open in a dim environment; in the DS-MWT, participants perform a monotonous, nighttime car-following task in a driving simulator. After each session, participants answer the KSS questionnaire again.

All sessions include continuous EEG monitoring and video recording. ET is implemented in all DS-MWT sessions and in MWT sessions where instrumentation is available. Following each run, bio-samples — including blood, saliva, exhaled breath, and dried blood spots — are collected.

The presence time for visits 3, 4, 6, and 7 is estimated at 8 hours each, including approximately 4 hours of active testing (4 × 40-minute sessions) and 15-minute post-session sampling intervals. EEG electrodes are removed after visits 4 and 7, but are checked for signal quality during all relevant visits and adjusted if needed.

Participants are invited to provide feedback upon study completion, either via an online form or in a personal follow-up conversation. An optional post-study interview is available for participants who wish to discuss their experiences, perceived effects, or reflections on the intervention’s impact on daily life.

### Randomization

Participants will be randomized to one of two starting conditions—either CPAP withdrawal (W) or continuation (C)—and assigned to one of two test sequences (*D → M* or *M → D*), which determine the measurement sequence. Randomization will occur following successful screening and is implemented via random lot drawing from an opaque lottery box containing sealed paper assignments. The randomization scheme can be seen on Fig.2

**Fig 2.**
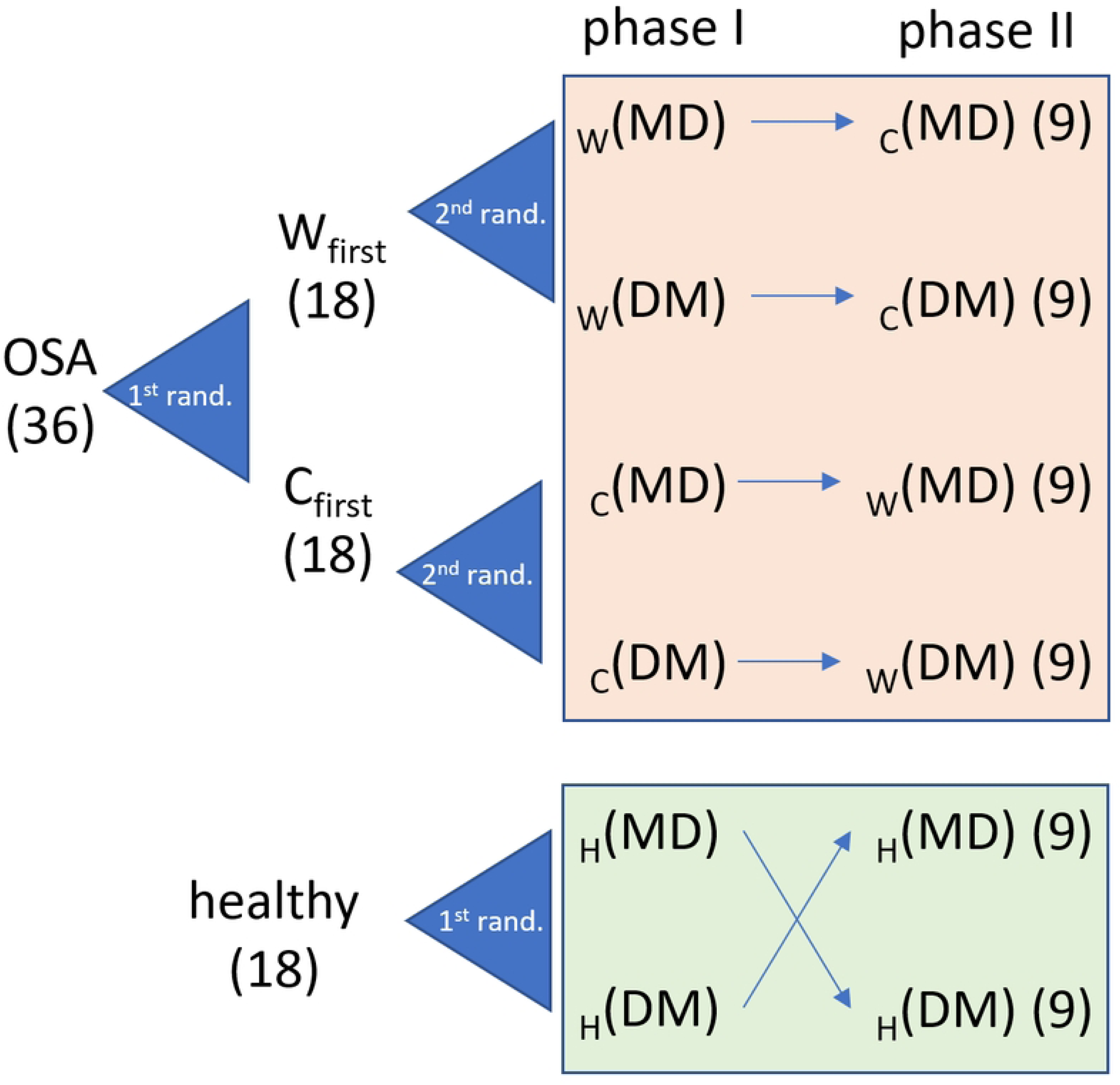
Overall randomization scheme for the intervention group and the comparison group. W_first_ = Group starting with the sequence in the condition “withdrawal”, i.e. the intervention. C_first_ = Group starting with the sequence in the condition “continuation”. D = driving simulation-based maintenance of wakefulness test, M = maintenance of wakefulness test.

No stratification will be applied based on demographic or clinical characteristics (e.g., sex, age, medication, dosage, or treatment duration). However, exploratory post hoc analyses will assess whether these factors influence outcome measures.

To ensure allocation concealment, the randomization lots are prepared in advance by the sponsor-investigator, who is not involved in the enrollment or assignment process. This separation preserves the integrity of the randomization and minimizes selection bias.

Due to the nature of the intervention, full blinding of participants and investigators is not feasible. Therefore, no formal unblinding procedure is necessary. However, outcome assessors will remain blinded to group allocation, including post-test, in order to prevent bias in the evaluation of subsequent participants.

### Outcomes

The primary outcome measure of this trial will be the difference in mean latency between the classical MWT and the DS-MWT. Sleep latency is determined according to the American Academy of Sleep Medicine (AASM) criteria [26], and DS-MWT latency is defined by the occurrence of the sleep accident (SA) with eyes closed during driving [5]. For both MWT and DS-MWT, the mean latency will be calculated as the average of all latencies across four standardized test sessions (each lasting a maximum of 40 minutes) conducted on the same test day.

For the secondary outcomes, the trial will include several measurement variables and assessments, designed to evaluate a broader range of physiological, behavioral, and subjective markers of sleepiness and driving performance.

1. Subjective sleepiness assessments:

- Situational Sleepiness: Participants will complete the Karolinska Sleepiness Scale (KSS) questionnaire immediately before and after each test run (at 9 a.m., 11 a.m., 1 p.m., and 3 p.m.) to assess momentary fluctuations in alertness [27, 28].
- General Daytime Sleepiness: At the screening visit, participants will complete the Epworth Sleepiness Scale (ESS) to establish a baseline measure of habitual EDS [29].
2. Driving performance metrics: The investigators will analyze whether driving behavior parameters, including SDLP, standard deviation of velocity (SDS), distance to the leading vehicle in the car-following task (SDD), and steering wheel angle (SDSW), correlate with CPAP treatment status and subjective sleepiness scores.
3. 3. EEG-based markers of sleepiness: The investigators will examine electrophysiological correlates of EDS, including MSEs and individual EEG features of the onset and transition states of sleep [30], and their association with CPAP withdrawal and subjective sleepiness (ESS, KSS).
4. Eye-tracking correlates: The investigators will examine whether eye-tracking features (e.g., blink rate, blink duration, eye openness, gaze direction, and attention to defined areas of interest) reflect treatment condition and align with subjective and objective markers of sleepiness
5. Night-to-night variability in sleep architecture: The investigators will assess sleep quality parameters and variability in OSA-related PSG parameters (e.g., AHI, ODI, mean SpO_2_, time *<* 90% saturation [*t<* 90], arousal-index) across repeated at-home-PSG measurements. Correlations with behavioral and subjective outcomes will be analyzed, and CPAP device data will be used to monitor treatment adherence [24].
6. Biomarker profiling: The investigators will evaluate whether previously identified proteomic and metabolomic biomarkers of sleepiness in non-invasive samples—such as saliva, exhaled breath, and dried blood spots [17] —correlate with CPAP status and subjective sleepiness ratings.

### Data collection and management

#### Data collection

All data is recorded semi-automatically (driving data, EEG, ET) and/or without interference from the investigator. To maximize both temporal accuracy and data reliability, the study employs a dual-recording strategy. First, all multimodal data ET, EEG, and driving simulation output (SILAB)—are recorded and synchronized in real time using the Lab Streaming Layer (LSL) framework [31]. This ensures precise temporal alignment across systems during active data collection. Concurrently, each subsystem independently records high-resolution raw data in its native format, providing a redundant backup to safeguard against potential streaming failures. This local storage approach guarantees data completeness.

To promote the quality and integrity of the collected data, the study team is trained in standardized procedures for assessment and documentation. The course of data collection is documented using standardized paper-based Case Report Forms (CRFs), completed by study investigators for each participant. These forms record all trial-relevant tests and assessments. Participants are identified by a unique participant ID (PID), which is assigned sequentially. A separate subject identification log links PIDs to participant identities and tracks screening outcomes, withdrawals, and dropouts.

All entries into the CRFs are verified against the source documents, which include medical records, laboratory results, psychological assessments, and other original documentation. In cases where data are directly extracted from instruments and not transcribed into the CRF—such as EEG or eye-tracking recordings—these are clearly identified as source data. The specific location of such source data is documented in the CRF in the designated data storage log.

#### Trial Instruments and Measures

For the primary outcome, EEG recordings are required to determine MWT-latencies, based on precise identification of sleep onset (SO). According to AASM classification, SO is defined as the start of the first epoch scored as any stage other than stage W (wake). In most individuals, this will usually be the first epoch of stage N1, characterized by SEM (slow eye movements) and LAMF (low-amplitude, mixed-frequency) in EEG activity [26]. For the calculation of DS-MWT-derived latencies, the classical EEG-based SO is complemented by the capture of SA in a driving simulator [5].

As for secondary outcome measures:

1. In addition to determining primary sleep latency outcomes, electrophysiological data collected during both MWT and DS-MWT sessions will provide insights into the dynamics of the sleep-wake transition. Of particular interest are MSEs —brief intrusions of sleep-like EEG activity during otherwise wakeful states —which may precede or occur independently of formal SO. Automatically detected MSEs will be analyzed in this study to characterize their distribution, duration, and relationship to subjective and behavioral sleepiness [30]. All EEG recordings, accompanied with electrooculogram (EOG), electromyogram (EMG), and electrocardiogram (ECG) —including those conducted during in-laboratory test sessions and overnight at-home PSG —will be performed using the Nox A1 PSG System (Nox Medical, Reykjavík, Iceland). This device has been validated for clinical sleep research and supports high-fidelity multi-channel EEG acquisition in both ambulatory and controlled lab environments [32]. Recordings will be analyzed using the dedicated Noxturnal software (Noxturnal, Nox Medical, Reykjavík, Iceland) and custom analysis scripts implemented in MATLAB (Version R2023a, The MathWorks, Inc., Natick, MA, USA).
2. Driving performance will be assessed using a custom-modified full-chassis BMW i3 simulator, engineered to deliver a high degree of ecological validity. The simulator is equipped with actuator systems replacing standard shock absorbers to simulate realistic motion cues, including acceleration, deceleration, road texture, and cornering forces. The vehicle is embedded within a 270-degree projection system offering 360-degree field-of-view coverage, thereby immersing participants in a virtual driving environment that closely mimics real-world conditions. The simulated environment is powered by SILAB^®^ Driving Simulation Software (Scenario Package “Driver Fitness and Ability,” Version 7.0; Würzburg Institute for Traffic Sciences GmbH, Veitshöchheim, Germany), which delivers a suite of validated scenario modules for evaluating FTD [33]. SILAB records a broad set of vehicle dynamics and behavioral parameters, including SDLP, SDS, number of lane departures, etc. Participants will perform a customized driving task, which consists of an approximately 40-minute, highly monotonous car-following task in a simulated nighttime setting, chosen to increase the likelihood of drowsiness-related events. The scenario simulates a low-stimulation, low-traffic environment, requiring sustained attention and minimal steering correction, closely mimicking real-world conditions where sleep-related driving incidents are most likely to occur.
3. Subjective sleepiness will be evaluated using computer-based questionnaires (KSS, ESS), administered and scored according to validated standard procedures.
4. Eye-tracking metrics will be continuously recorded during DS-MWT and MWT sessions using high-resolution eye-tracking systems Tobii Pro Glasses 3 (Tobii AB, Danderyd/Stockholm, Sweden). A range of ocular motor features, including blink rate, saccade latency, saccade peak velocity and smooth pursuit velocity gain, will be analyzed. These parameters will be examined to determine whether they reflect levels of sleepiness and whether they hold predictive value for SO or MSE during the driving simulation [34, 35].
5. Unstimulated oral fluid will be collected using neutral Salivette^®^ devices (Sarstedt, Sevelen, Germany), following standardized procedures for non-invasive saliva sampling [36]. Samples will be immediately frozen and stored at *−*80 *^◦^*C until the conclusion of the trial. After completion of data collection, samples will undergo targeted metabolomic analysis using ultra-performance liquid chromatography coupled to high-resolution mass spectrometry (UPLC-HRMS) [18]
6. Dried blood spot (DBS) samples will be collected using MICROLET^®^ lancets (Bayer HealthCare LLC, Whippany, NJ, USA) applied onto DBS collection cards (CAMAG AG, Muttenz, Switzerland), which allow for precise, standardized blood volume collection. All DBS samples will be stored under controlled conditions and subsequently processed and analyzed analogously to oral fluid samples.
7. Exhaled breath samples will be collected using MIR Spirobank Smart spirometer (MIR, Rome, Italy) and the BreathExplor^®^ device (Munkplast AB, Uppsala, Sweden). These devices are designed for safe, efficient collection of aerosol particles from the exhaled air, suitable for subsequent biochemical and metabolomic analyses.

#### Data Management

All aspects of data management are coordinated by the Division of Traffic Medicine, which also serves as the central coordinating body for the study. Data are handled in compliance with Swiss data protection regulations. CRFs and other study documents are stored in locked areas with access limited to authorized personnel. Biological samples are identified solely by PID, stored under secure conditions, and transported personally by authorized staff to the designated analysis laboratory with authorized restricted access. No genetic data are collected, and no biobank will be established.

In case of participant’s discontinuation, data and samples collected up to the point of withdrawal may be retained and analyzed. If a participant leaves early, no additional final visit is planned, but support is provided for a safe departure if needed.

After study completion, all study documents and data are archived for 20 years at the Institute for Forensic Medicine, University of Zurich. This includes CRFs, consent forms, medical records, and laboratory results. Data analysis will be performed only after trial completion and independently of funding sources. Data will be coded, processed, and analyzed in accordance with the predefined statistical analysis plan.

### Statistical methods

There is no dedicated statistician other than the investigators involved. However, a statistician will be available throughout the study and subsequent analyzes.

#### Sample size calculation

The total sample size is N_total_ = 54, consisting of the intervention group N_I_ = 36 and a healthy comparison group N_H_ = 18.

Sample size of N_I_ = 36 for the intervention group was estimated according Formula (1), taking into account the adapted randomized crossover RCT design [37]. The study involves a cohort of OSA patients randomized to both the CPAP starting condition and the measurement sequence, resulting in four groups (W-DM, W-MD, C-DM, C-MD) across two phases. A mixed-model ANOVA will be used for data analysis, as it appropriately accounts for the correlation of repeated measures within individuals, handles missing data more effectively, and incorporates both fixed and random effects.

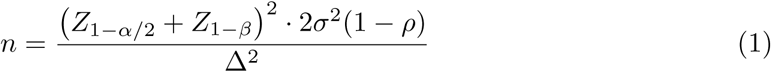

Where,

- *n* = Required number of participants per group.
- *Z*_1_*_−α/_*_2_ = Z-value corresponding to the chosen significance level (e.g., 1.645 for *α* = 0.1 two-sided) [38].
- *Z*_1_*_−β_* = Z-value corresponding to the desired power (e.g., 1.28 for 90% power) [38].
- *σ* = Standard deviation of the measurements = 13.5 minutes (from Bonnet & Arand [15]).
- *ρ* = Intra-class correlation (ICC), i.e., the correlation between repeated measures within the same participant.
- Δ = Equivalence margin or effect size = 1 pooled SD = 13.5 minutes (from Bonnet & Arand [15]).

For the sample size calculations, a two-sided *α* = 0.1 was used, assuming that a potential difference between M and D would have a clear direction and a maximal value (bounded test range) of 40 min for the test latency. When conducting an equivalence test, using a 0.1 two-sided alpha-level effectively matches the 0.05 significance level for each side of the distribution (one for each tail), aligning the rigor of the test with the traditional hypothesis test for no difference. This assumption holds, as the reference standard deviation of 13.5 min was obtained with an equivalently bounded test range. A graphically extracted and pooled standard deviation from Bonnet was used & Arand [15], the only close-to test-retest data on MWTs available.

Similarly, a sample size of N_H_ = 18 for the comparison group (healthy) was estimated for the comparison group of healthy participants, based on a crossover design without retest, as applied in the test-retest framework. This corresponds to 9 participants per group to achieve 90% power, assuming a two-way ANOVA (see Formula (2), adapted from Chow & Liu [39]), resulting in a total of 18 participants across both groups. Although a smaller standard deviation (*σ* = 5.9 minutes) was used — derived from normative data on healthy individuals reported by Banks et al. [12] — the required sample size remains comparable to that of the OSA cohort. This is primarily due to the absence of additional variability-reducing conditions that were present in the OSA sample.

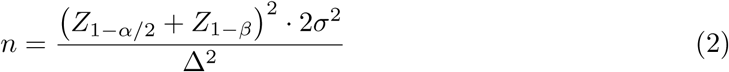

Where,

- *n* = Required number of participants per group.
- *Z*_1_*_−α/_*_2_ = Z-value corresponding to the chosen significance level (e.g., 1.645 for *α* = 0.1 two-sided) [38].
- *Z*_1_*_−β_* = Z-value corresponding to the desired power (e.g., 1.28 for 90% power) [38].
- *σ* = Standard deviation of the measurements = 13.5 minutes (from Bonnet & Arand [15]).
- *ρ* = Intra-class correlation (ICC), i.e., the correlation between repeated measures within the same participant.
- Δ = Equivalence margin or effect size = 1 pooled SD = 13.5 minutes (from Bonnet & Arand [15]).

#### Statistical analysis plan

Based on the design of this prospective, randomized, crossover trial, statistical analysis will be performed to detect both intra- and inter-individual (i.e., individual and group-level) differences. All analyses will be conducted in relation to two key contextual variables:

- Sleep quality of the night preceding each test session, as assessed by PSG, and
- Time of day at which each test run was performed (9 a.m., 11 a.m., 1 p.m., 3 p.m.).

The primary hypothesis will be tested solely based on the determined mean sleep latencies obtained from the MWT and DS-MWT, without incorporating additional secondary or exploratory parameters. For the primary outcome, a mixed model ANOVA will be applied or, alternatively, a generalized linear mixed model (GLMM). Additional covariates, such as prior night PSG outcomes and time of the day, will be included. Results will be considered significant if p *<* 0.05.

All secondary variables will be analyzed post hoc, with the goal of generating new hypotheses rather than performing formal confirmatory tests. These analyses will include individual evaluations of each collected secondary parameter in reference to:

- The experimental condition (MWT vs. DS-MWT),
- The treatment state (CPAP continuation vs. withdrawal) or healthy comparison,
- The test sequence (DM vs. MD).

Where appropriate, group-level comparisons will be followed by multiple comparison tests to examine differences in greater detail. For example, time-course effects in driving behavior parameters—such as SDLP —will be examined using repeated measures ANOVA across ten 4-minute segments within each test session, or through paired t-tests comparing the first and final modules of each session. Additionally, Integrated Driving Scores (IDS) will be computed as Z-scores following logistic regression models derived from driving performance metrics.

For these secondary analyses, linear mixed models will be used as the initial framework, with stepwise model testing based on biologically plausible relationships rather than relying solely on statistical significance. Where applicable, Akaike Information Criterion (AIC) will be used to identify the most parsimonious model, and AICc will be applied in cases involving small sample sizes (N=18). Cross-validation techniques, such as k-fold cross-validation, will be implemented to evaluate predictive performance and reduce overfitting.

Beyond aggregated latency measures, predictive models for SO and/or SA will be explored at finer temporal resolutions, including:

- Per 40-minute run (N = 4 runs × 54 participants = 216 observations),
- Per 4-minute driving module (N = 10 modules × 216 runs = 2,160 observations),
- Or via dynamic time-series analysis using shorter time units still to be defined.

These exploratory analyses will aim to maintain an events-per-variable (EPV) ratio between 10 and 15, depending on the number of predictors included. These models will evaluate whether secondary variables collected during each run can predict the occurrence of SO or MSE in real time.

While p-values will not be the primary decision metric for exploratory models, False Discovery Rate (FDR) corrections may be applied to account for multiple testing while preserving statistical power. Bonferroni corrections, due to their conservativeness, will be avoided to reduce the risk of Type II errors (false negatives). Significant findings from exploratory analyses will be interpreted cautiously and treated as hypothesis-generating, requiring independent replication in future studies.

Although our study is not powered to detect gender differences, sex/gender will be included as a covariate in all relevant models for control purposes.

All statistical analyses will be performed using R, Python, MatLab and GraphPad Prism. Any deviations from this predefined statistical analysis plan will be documented and justified in the final trial report.

The study is intended to be completed with N = 54 participants, unless recruitment proves infeasible within the projected time frame. The last participant’s last visit (LPLV) is planned for 30 April 2027, though an extension of the recruitment period may be considered if necessary.

Missing datasets due to participants’ withdrawal from the trial will not be multiply imputed due to the low sample size and inherently low statistical power. All available data will be used, if possible, for statistical analysis. Drop-outs during the study phase will be replaced by recruitment of new participants from the accessible participant pool, if possible.

## Discussion

The classical maintenance of wakefulness test (MWT) is a widely accepted method for assessing EDS across various clinical conditions, although its broader applicability remains debated [4]. The MWT evaluates the ability to remain awake in a quiet, non-stimulating environment [40]. It is often used to assess the presence of EDS, to monitor treatment response in sleep disorders, and to evaluate wakefulness in individuals whose sustained alertness is safety-critical [14]. While valuable in this context, the MWT may not fully capture the challenges of driving, which involves cognitive engagement and environmental stimulation. This protocol seeks to establish a more ecologically valid proxy for real-world driving risks than conventional MWT. It remains to be examined whether DS-MWT latencies differ significantly from MWT latencies. Such a finding would support the view that traditional MWT alone may under- or overestimate real-life driving impairment in sleepy individuals. This could inform clinical and regulatory practice regarding FTD assessments in patients with EDS or sleep disorders, potentially leading to a meaningful refinement in how sleepiness-related driving risk is evaluated in traffic medicine.

By embedding the wakefulness assessment in a realistic driving simulation — a monotonous, nighttime, car-following task — the driving simulation-based maintenance of wakefulness test (DS-MWT) may capture subtle impairments in vigilance, attention maintenance, and vehicle control that traditional MWT, lacking behavioral and environmental context, might miss.

Moreover, the extended set of secondary outcomes, including driving behavior metrics (e.g., SDLP, SDS, lane departures, etc.), eye-tracking and EEG parameters, subjective sleepiness ratings, polysomnographic sleep quality data, and biomarker analyzes (saliva, exhaled breath, and DBS), offers the possibility for a multimodal characterization of sleepiness and alertness. If associations emerge among these parameters in this naturalistic test setting, they may suggest objective, sensitive markers of sleepiness that better predict risk of impaired driving than classical latency measures alone. Ultimately, this could support the development of novel, non-invasive screening tools for real-world drowsy driving risk, benefiting public health and potentially reducing preventable traffic accidents. In addition, by using a within-subject crossover design with CPAP withdrawal and resumption, the study may yield insights into the dynamics of EDS and its reversibility under therapeutic intervention. This could reinforce the importance of CPAP adherence in OSA management.

## Limitations

A major strength of the design is the within-subject crossover format, which controls for inter-individual variability in sleep physiology, OSA severity, medication status, and other confounders. Inclusion of a well-adherent OSA patient pool, pre-screened for comorbidities, ensures clinical safety and homogeneity. However, several limitations are clearly present. First, blinding is not feasible for participants or study personnel due to the nature of the intervention (CPAP withdrawal and simulation vs. classical MWT). Although outcome assessors will be blinded where possible and data acquisition is automated, some bias cannot be excluded — for example, participants’ awareness of driving simulation might influence their subjective sleepiness ratings, motivation, or compensation behaviors. This may limit the extent to which DS-MWT findings can be generalized to unmonitored, real-life driving situations where motivation, distraction, or compensatory behaviors differ. Second, the multiplicity of secondary and exploratory analyses increases the risk of false-positive findings or overfitting if included in one model. While the planned use of model-selection criteria (AIC/AICc), cross-validation, and FDR correction mitigates this risk, such methods do not fully eliminate it — and any promising associations will require independent replication in larger, prospective cohorts. Therefore, the results of this study can provide a foundation for further studies evaluating the DS-MWT in clinically and demographically diverse populations — including CPAP-naive individuals, patients with other sleep disorders, shift workers, and professional drivers. Moreover, if certain biomarkers or eye-tracking / EEG features prove to correlate robustly with simulated driving impairment, future work could aim to develop non-invasive screening tools for use in clinical, occupational, or roadside screening settings. This could be complemented by machine-learning approaches trained on multi-modal data to predict drowsiness or microsleep risk in individual drivers. In conclusion, this trial might bridge existent gaps in sleep and traffic medicine by evaluating a novel, ecologically valid simulation-based assessment of wakefulness and driving performance in OSA patients and healthy controls. In conclusion, by combining classical neurophysiological measures, behavioral simulation, subjective sleepiness scales, and multimodal biomarker sampling, the study has the potential to significantly advance our understanding of how EDS translates into driving risks.

## Data Availability

No datasets were generated or analysed during the current study. All relevant data from this study will be made available upon study completion.

## Abbreviations

AASM: American Academy of Sleep Medicine
AE: Adverse Event
AIC: Akaike Information Criterion
ASR: Annual Safety Report
C CPAP: continuation
ClinO: Ordinance on Clinical Trials in Human Research (in German: KlinV, in French: OClin, in Italian: OSRUm)
CPAP: continuous positive airway pressure
CRF: Case Report Form
D, DS-MWT: driving simulated maintenance of wakefulness test
DS: driving simulation
EDS: excessive daytime sleepiness
EEG: Electroencephalogram
ESS: Epworth Sleepiness scale
ET: Eye-tracking
GCP: Good Clinical Practice
HRA: Human Research Act (in German: HFG, in French: LRH, in Italian: LRUm)
ICH: International Council for Harmonisation
IDS: integrated driving score
KSS: Karolinska Sleepiness scale
M, MWT: maintenance of wakefulness test
OSA: Obstructive sleep apnea
PID: unique participant
ID SA: Sleep Accident
SDD: distance to the leading vehicle in the car-following task
SDLP: tandard deviation of lane position
SDS: standard deviation of velocity
SDSW: steering wheel angle
V: Visit
V: CPAP withdrawal

## Administrative information

### Monitoring

Internal monitoring duties will be carried out by Dr. med. Kristina Keller, who is not actively taking part in recruitment, data acquisition or data analysis. There is no additional external monitoring foreseen or required. Internal monitoring will be conducted to ensure adherence to the study protocol, regulatory requirements, and data quality standards. All source data and relevant documents will be accessible to the internal monitor. Monitoring visits may take place without prior notice and will include a review of available documentation and clarification of any issues directly with the site team.

Given the exploratory nature of this study, the establishment of a steering committee, an endpoint adjudication committee, or an independent data monitoring committee is not required and has therefore not been implemented.

### Ethics

#### Research Ethics Approval

This study is conducted in accordance with the current version of the Declaration of Helsinki, the International Council for Harmonisation Good Clinical Practice (ICH-GCP) guidelines, the Human Research Act (HRA), and all other locally applicable regulatory requirements. The trial protocol and related study materials were reviewed and approved by the Kantonale Ethikkommission Zürich (Zurich Cantonal Ethics Committee), with the reference number 2024-01948. No participant recruitment or data collection activities commenced prior to obtaining this ethics approval.

#### Protocol Amendments

Any substantial amendments to the protocol, including changes to study objectives, design, methodology, study site(s), sponsor, or key study personnel, will be submitted for review and approval by the Ethics Committee prior to implementation. Exceptions may only occur under emergency circumstances to ensure participant safety and well-being; such deviations shall be documented and reported to the Ethics Committee as soon as possible.

Substantial amendments are defined in accordance with ClinO (Ordinance on Clinical Trials in Human Research), Art. 29, and include changes affecting participant safety, rights, or obligations. A list of substantial changes is maintained on www.swissethics.ch. Non-substantial amendments will be compiled and submitted annually to the Ethics Committee together with the Annual Safety Report (ASR).

#### Confidentiality

Participant confidentiality will be maintained throughout the trial in compliance with Swiss data protection regulations. Identifiable information is stored separately in a subject identification log, linking names to PIDs assigned in sequence of enrolment.

This log also records reasons for exclusion, early withdrawal, or non-adherence.

Access to identifiable data is restricted to authorized personnel involved in the study. Any disclosure to third parties requires prior written permission from the data owner. Study data may be accessed by monitors or the Ethics Committee solely for verification purposes.

#### Harms

This study is classified as category A, involving only minimal risk to participants. The procedures conducted across the seven study visits consist predominantly of standardized, non-invasive assessments, performed under continuous supervision by trained research personnel.

The use of a driving simulator may occasionally cause dizziness, nausea, or discomfort due to simulator sickness. These symptoms typically subside promptly upon test completion or leaving the simulator environment.

EEG monitoring and blood collection via a peripheral venous catheter are performed by qualified personnel. Minor adverse events, such as hematoma at the venipuncture site or skin irritation from EEG electrodes, may occur but are generally transient and resolve without complications. The total blood volume collected (9 mL) poses no significant health risk. The collection of saliva, urine, DBSs, and exhaled breath is entirely non-invasive and carries no known risks.

Participants assigned to the intervention arm will discontinue their nightly CPAP therapy at home for a period of *≥*7 days, during which a temporary reactivation of OSA symptoms is expected. This CPAP-withdrawal model is clinically validated as a safe and effective method for studying the pathophysiology of OSA [22, 41]. Only patients deemed low risk for adverse outcomes by physicians at the Sleep Disorders Center, based on clinical judgment and prior screening, will be included. Participants will resume their established long-term CPAP therapy immediately after the study.

All procedures will be conducted in controlled environments by trained staff, with medical oversight provided as needed. Participants are closely monitored for any adverse effects, and appropriate support will be made available. Individuals retain the right to withdraw from the study at any point without consequences for their ongoing medical care.

Participation in the study offers no direct therapeutic benefit to the individuals involved. However, the findings may contribute to the advancement of diagnostic and safety tools in traffic medicine and sleep disorder management, potentially benefiting future patients and public health initiatives.

### Roles and responsibilities

SL is the sponsor and principal investigator of the project; he developed the design of the trial in collaboration with EIS. SL acquired the funding for the trial and drafted the study proposal for ethics submission. The funding body did not contribute to the study design and will not be involved in its execution, data analysis, interpretation, or the decision to publish the results. VG is responsible for trial management; she contributed to the design and development of the protocol, as well as the drafting of the manuscript. NL contributed to the design and development of the protocol. EIS is a co-investigator and recruitment partner. KK is the study site leader and the investigator physician. All authors read and approved the final manuscript.

## Data availability

Trial and participant data will be handled with uttermost discretion and are only accessible to authorized personnel who require the data to fulfill their duties within the scope of the study. On the CRFs and other study-specific documents, participants are only identified by a unique participant number.

Any data necessary to support the protocol can be provided upon request. Statistical code will be made publicly available on the GitHub repository after the publication of results. The SPIRIT checklist can be found in the Supporting information chapter.

The results of this study will be published in peer-reviewed scientific journals, regardless of the direction of outcomes. Digital copies will be sent to trial participants if they express their interest. Findings will also be presented at scientific conferences and meetings. A final project report will be sent to the funder.

## Supporting information

**S1 File. SPIRIT 2025 checklist.**

**S2 File. DS-MWT2 study protocol approved by the ethics committee.**

## Acknowledgments

We thank Lana Brockbals and Michael Scholz (Forensic Pharmacology & Toxicology, Institute of Forensic Medicine, University of Zurich) for the discussion on sample taking and future analyses.

